# Scoping review of factors associated with stem cell mobilisation and collection in allogeneic stem cell donors

**DOI:** 10.1101/2024.03.15.24304360

**Authors:** Rachel C Peck, Amber Knapp-Wilson, Kate Burley, Carolyn Dorée, James Griffin, Andrew D Mumford, Simon Stanworth, Kirsty Sharplin

## Abstract

**Background:** There is a large inter-individual variation in CD34+ cell yield after G-CSF mobilisation and collection from peripheral blood in healthy allogenic haematopoietic stem cell donors. Donor characteristics including gender and age, baseline and pre-collection blood results, mobilisation factors and collection factors have been associated with CD34+ cell concentration in the blood after G-CSF mobilisation and/or CD34+ cell yield after collection. Since the literature reporting these associations is heterogeneous, we here clarify the determinants of CD34+ cell concentration and yield through a scoping literature review.

**Materials and Methods:** MEDLINE, Embase, PubMed and Stem Cell Evidence were searched for studies published between 2000 and 2023. The inclusion criteria were studies of allogeneic donors undergoing G-CSF mobilisation and peripheral blood stem cell collection (PBSC). Eligible studies assessed an outcome of mobilisation or collection efficacy, indicated by the blood CD34+ cell concentration after 4 or 5 days of G-CSF treatment and/or CD34^+^ cell yield in the first PBSC collection after mobilisation. Included studies assessed associations between these outcomes and donor factors (such as age, gender, weight, ethnicity), mobilisation factors (G-CSF scheduling or dose), collection factors (venous access, processed blood volume) and laboratory factors (such as blood cell counts at baseline and after mobilisation).

**Results:** The 51 eligible studies evaluated between 23 and 20,884 donors. 43 studies were retrospective, 32 assessed blood CD34+ cell concentration after mobilisation and 37 assessed CD34+ cell yield. In studies that recorded both outcomes, blood CD34+ cell concentration always predicted CD34+ cell yield. The most frequently assessed factor was donor age for which most studies reported that younger donors had a higher blood CD34+ cell concentration and CD34+ cell yield. Non-European ancestry was associated with both higher blood CD34+ cell concentration and yield although this finding was inconsistent.

**Conclusions:** There remains poor consensus about the best predictors of blood CD34+ cell concentration and yield that requires further prospective study, particularly of the role of donor ancestry. The current focus on donor gender as a major predictor may require re-evaluation.

## INTRODUCTION

Allogeneic haematopoietic stem cell transplantation (HSCT) is a life-saving treatment for haematological and solid-organ malignancies and most commonly uses stem cells (CD34^+^ cells) from peripheral blood obtained through a two-stage process: i. Mobilisation of CD34^+^ cells from bone marrow using the cytokine G-CSF for 4-5 days, and ii. Peripheral blood stem cell collection (PBSC) by apheresis.

Despite standardised mobilisation and PBSC protocols, there is marked inter-individual variation in the number of CD34+ cells mobilised and collected (1, 2). Although most allogeneic donors achieve target CD34^+^ cell yield (typically 4-5 ×10^6^/kg CD34^+^ cells/recipient weight), only 60% do this with one PBSC. The remaining 40% of donors usually achieve target yield after repeat PBSC, although 2-5% are poor mobilisers and never achieve target. For recipients, suboptimal CD34^+^ cell yields from donors result in prolonged engraftment times and increased risk of adverse events such as graft failure, infection, or bleeding (3, 4).

Numerous donor characteristic and collection factors have been postulated to impact on successful CD34+ cell donation, these can be broadly divided into *donor*, *mobilisation, collection* and *laboratory* factors. However, the existing literature contains abundant inconclusive and sometimes contradictory findings, often resulting from limitations such as small sample sizes, retrospective cohort designs and heterogenous outcome definitions. The objective of this scoping review is to answer the research question *What are the donor, mobilisation, collection and laboratory factors that are associated with blood CD34^+^ cell concentration and/or CD34^+^ cell yield in allogenic donors after standard G-CSF mobilisation?* Standard G-CSF mobilisation is defined here as treatment of donors with short-acting recombinant human G-CSF for four or five-days duration. Although the CD34+ cell concentration and yield outcomes are anticipated to be closely correlated, they have been separated in this review as blood CD34^+^ cell concentration is potentially influenced only by donor and mobilisation factors whereas CD34+ cell yield may also be influenced by collection factors.

## MATERIALS AND METHODS

We conducted a scoping rather than a systematic review because of the breadth of previously investigated candidate predictors and because of the retrospective design and heterogeneity of most previous studies. A scoping review was anticipated to enable better mapping of key concepts and identification of knowledge gaps without the constraints imposed by the requirement of homogenous eligibility criteria needed for systematic review. The Preferred Reporting Items for Systematic reviews and Meta-Analyses extension for Scoping review (PRISMA-ScR) checklist was used to ensure best practice (5). The review protocol is available on request to the corresponding author.

### Search Strategy

Electronic databases (MEDLINE, Embase, PubMed and Stem Cell Evidence) were searched for studies published between January 1 2000 and November 1 2023 by a review author (CD) who is part of the Systematic Review Initiative team. The search strategy is documented in Supplementary Data.

### Eligibility criteria

Studies were eligible for analysis if they reported data from allogeneic stem cell donors who underwent G-CSF mobilisation and PBSC and if they reported at least one of *blood CD34+ cell concentration* after 4 or 5 days of G-CSF or *CD34^+^ cell yield* at the first PBSC. Studies were included if they evaluated associations between these outcomes and candidate variables in the following groups: i. *donor factors* (eg age, sex, weight, ethnicity); ii. *mobilisation factors* (eg G-CSF scheduling or dose); iii. PBSC *collection factors* (eg venous access, processed blood volume), or, iv. Donor *laboratory characteristics* at baseline or after mobilisation. Studies were excluded if they reported data from donors aged <16 years, or if they were case reports, reviews with no new data, non-English language publications, animal studies or pre-2000 studies.

### Study selection

After duplicate removal, two authors (RP and AKW) used the Rayyan© platform to screen titles and abstracts before resolving conflicts through discussion. Full article texts were then obtained for eligibility review by RP. The references of included studies and review articles were searched to ensure capture of all relevant studies.

### Data charting and analysis

Data was extracted into a standardised form which was piloted by and subsequently refined by RP and KB. The following data was extracted by RP: Authors, publication year, sample size, funding, country of study, donor demographic data, G-CSF protocol, make of apheresis machine, processed blood volume, anticoagulant, venous access, duration of collection, flow rate, and outcomes assessed.

## RESULTS

### Literature search

After duplicates were removed, 3355 articles were screened by the title and abstract. The remaining 118 articles were then screened by inspection of full text to yield 51 eligible studies for analysis (Figure 1). Studies were divided according to whether the reported outcome was only blood CD34^+^ cell concentration after G-CSF mobilisation (n = 14 Table 1), only CD34^+^ cell yield from first PBSC (n =18; Table 2) or if they reported both outcomes (n=19 listed in both Tables 1 and 2). Excluded studies are listed with explanatory notes in Supplementary Data.

**Figure 1:**
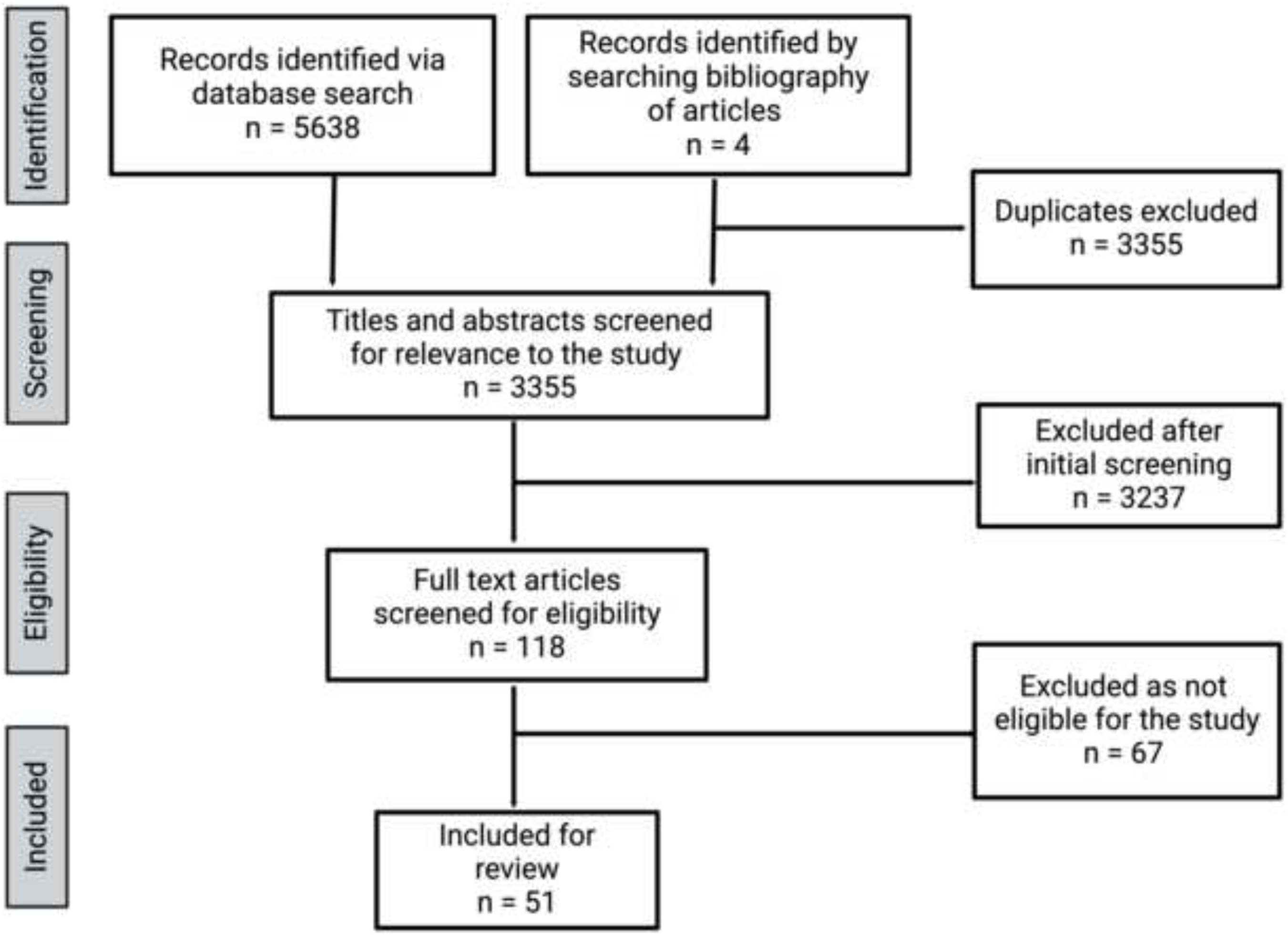
PRISMA Study flow chart.

**Figure 2:**
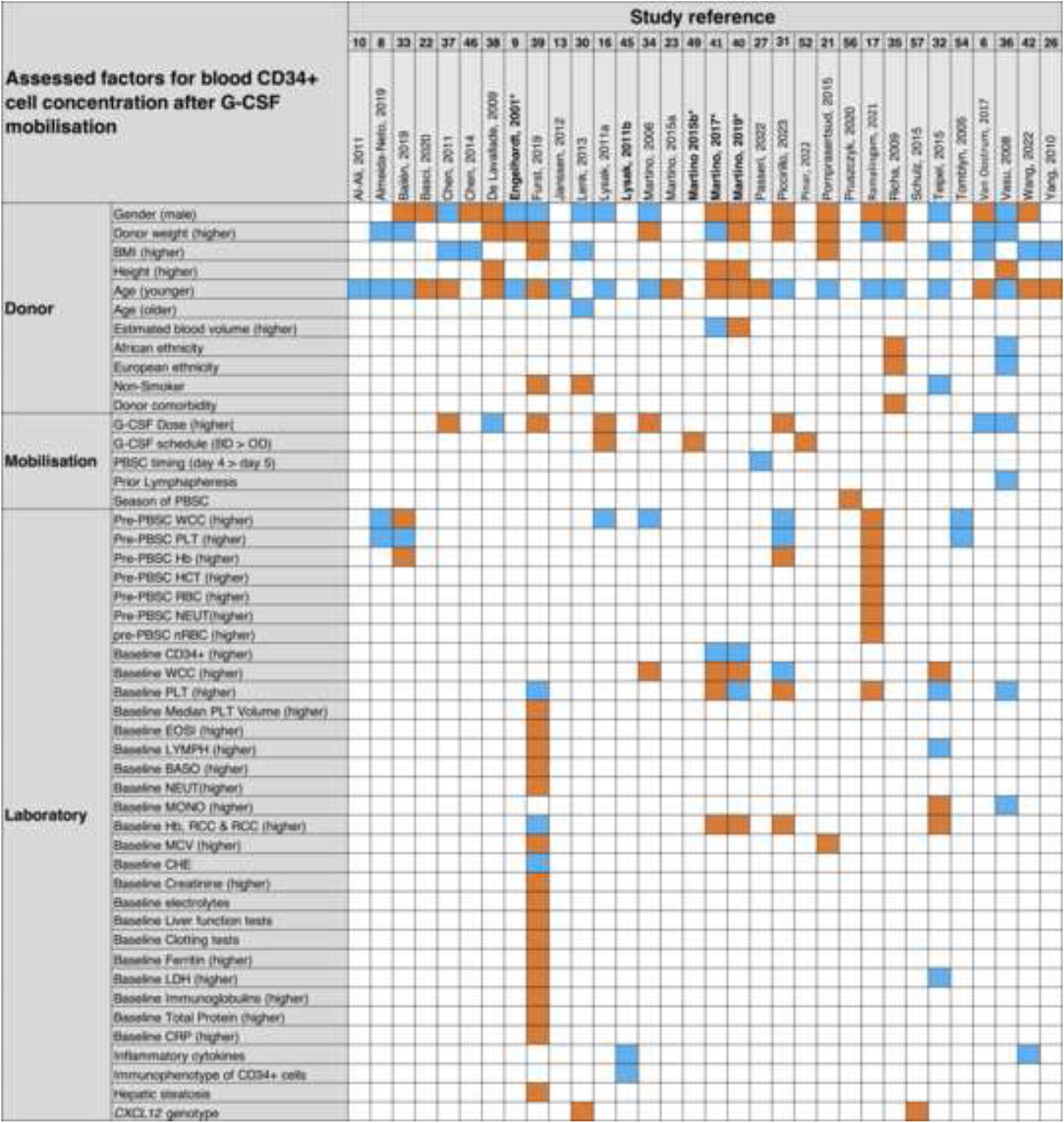
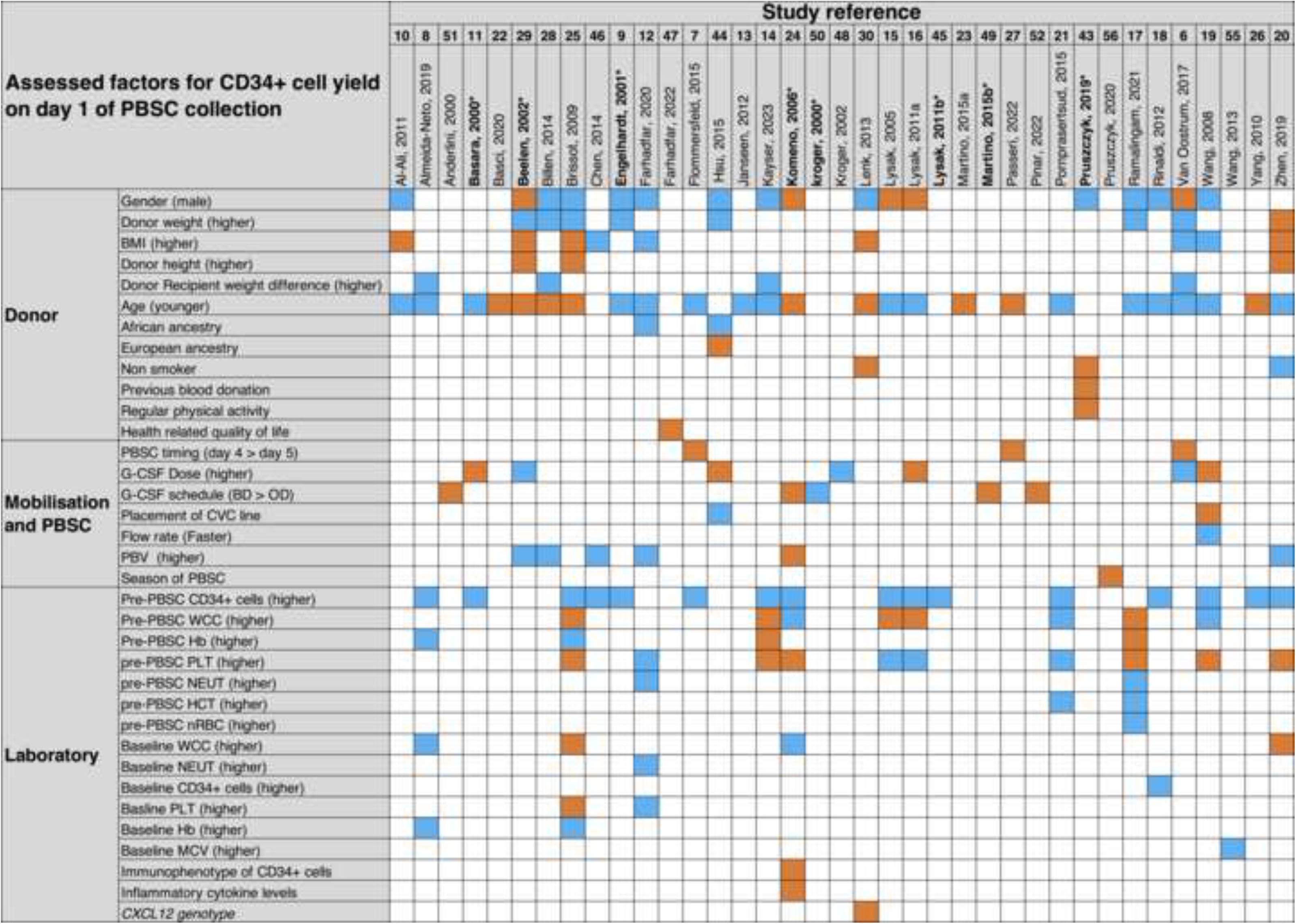
Heatmap of studies included in the scoping review. On the X axis are references of the studies in the scoping review. On the Y axis are the factors assessed by each study. Blue squares indicate a statistically significant association, orange squares indicate no statistically significant association. Figure 2A displays results of studies which assessed factors with the outcome of Blood CD34+ cell concentration after G-CSF mobilisation. Figure 2B displays results of studies which assessed factors with the outcome of CD34+ cell yield on day 1 of PBSC. References in **bold** indicate prospective or randomized control studies. BMI-Body Mass Index; PBSC-Peripheral blood stem cell collection; CVC-Central venous Catheter; WCC-White Cell count; Hb-Haemoglobin concentration; PLT-Platelet count; NEUT-Neutrophil count; HCT-Haematocrit; nRBC-nucleated red blood cell count; MCV-Mean Corpuscular Volume; MONO-Mononuclear cell count; EOSI-Eosinophil cell count; LYMP-lymphocyte count; CHE-Cholinesterase; electrolytes-Na, K, Ca, Mg2+; PBV-Processed Blood Volume

**Table 1:**
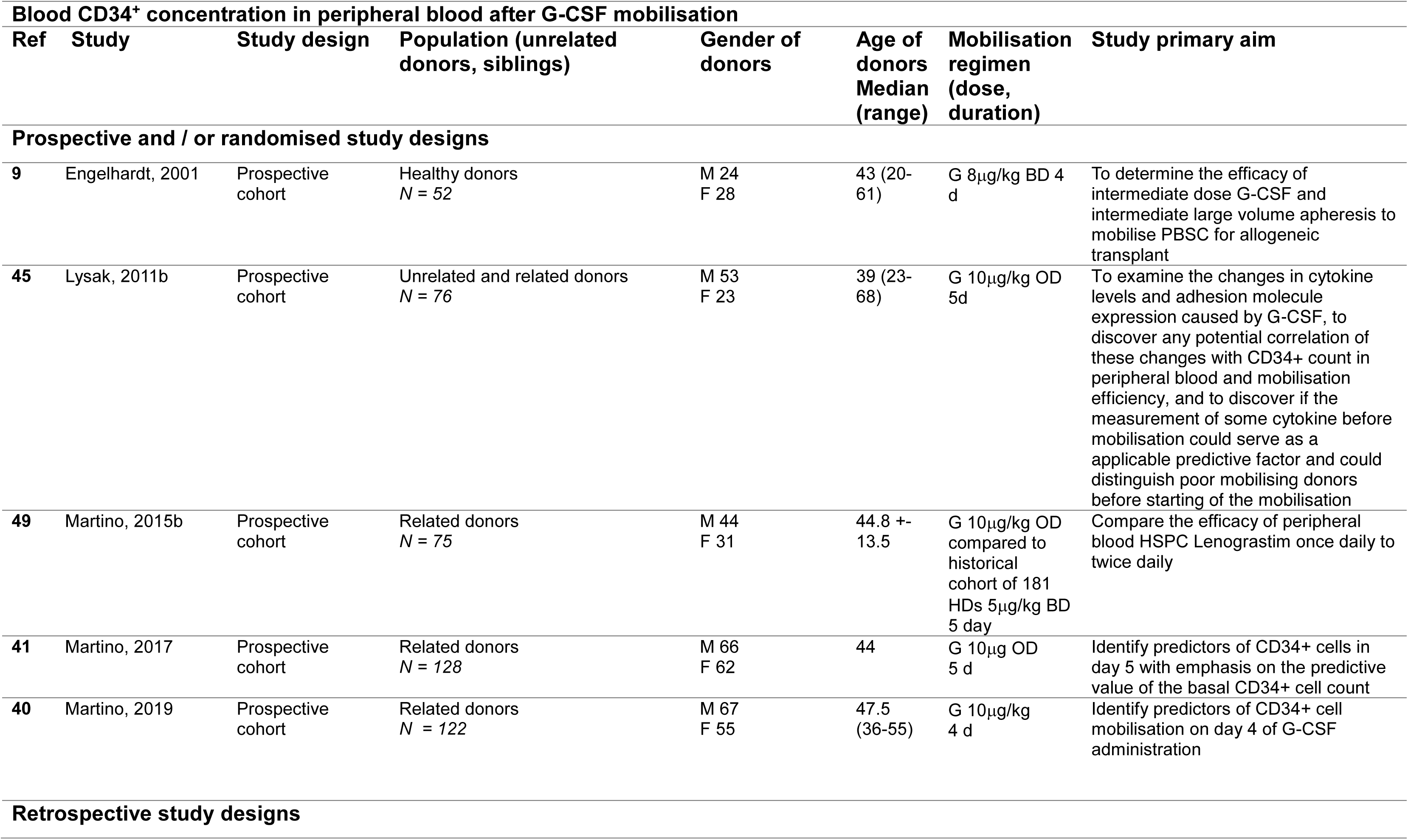
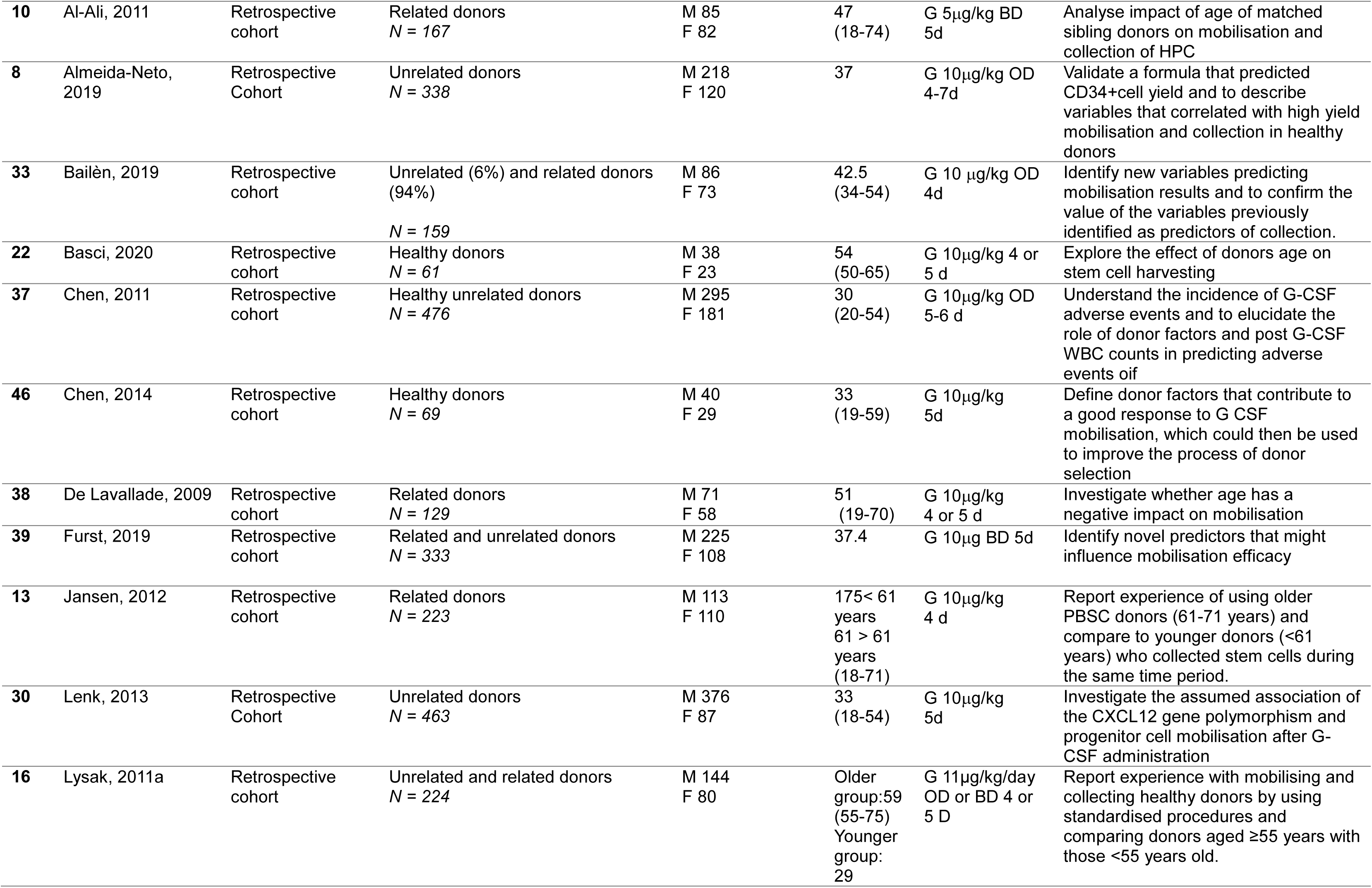
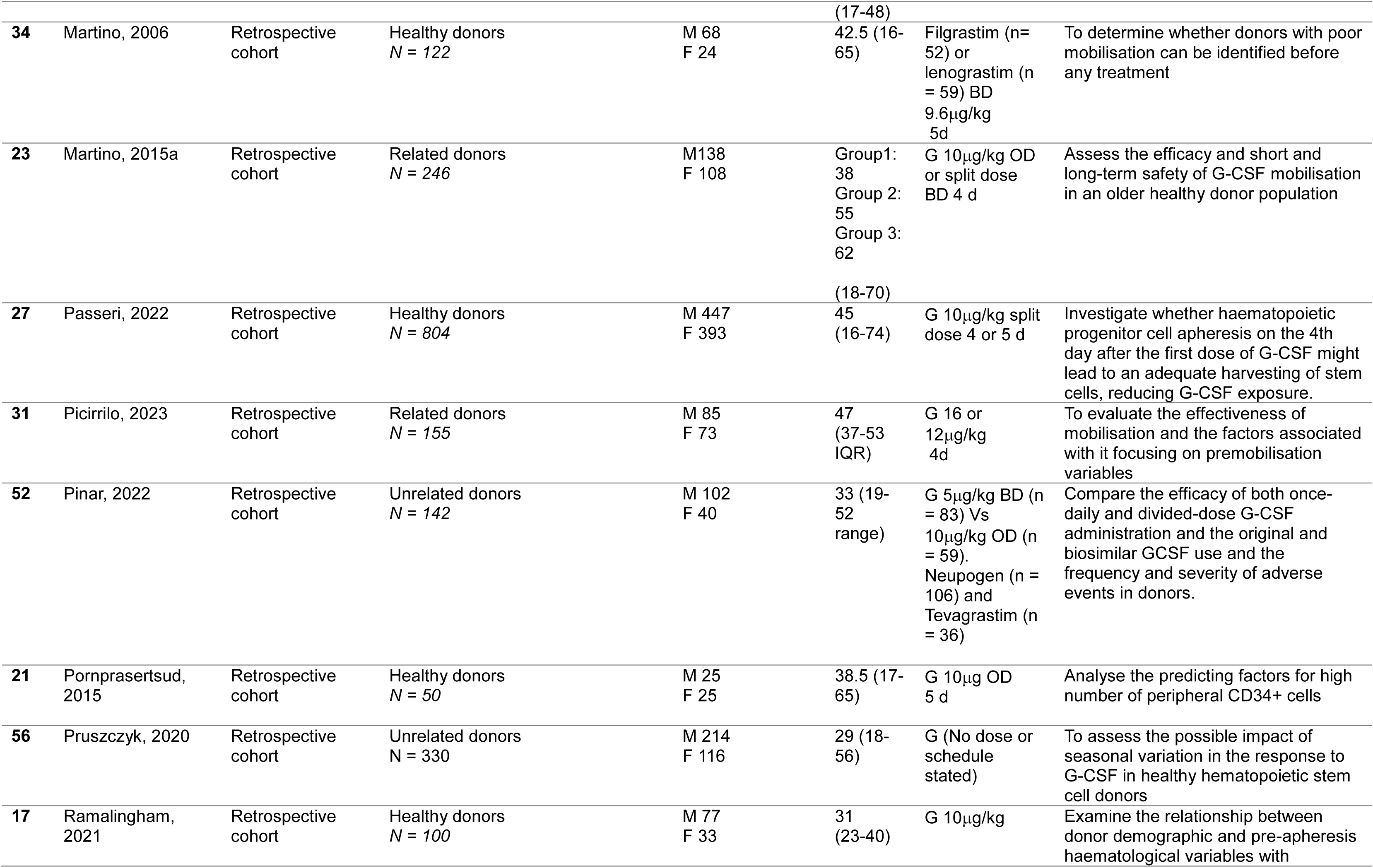
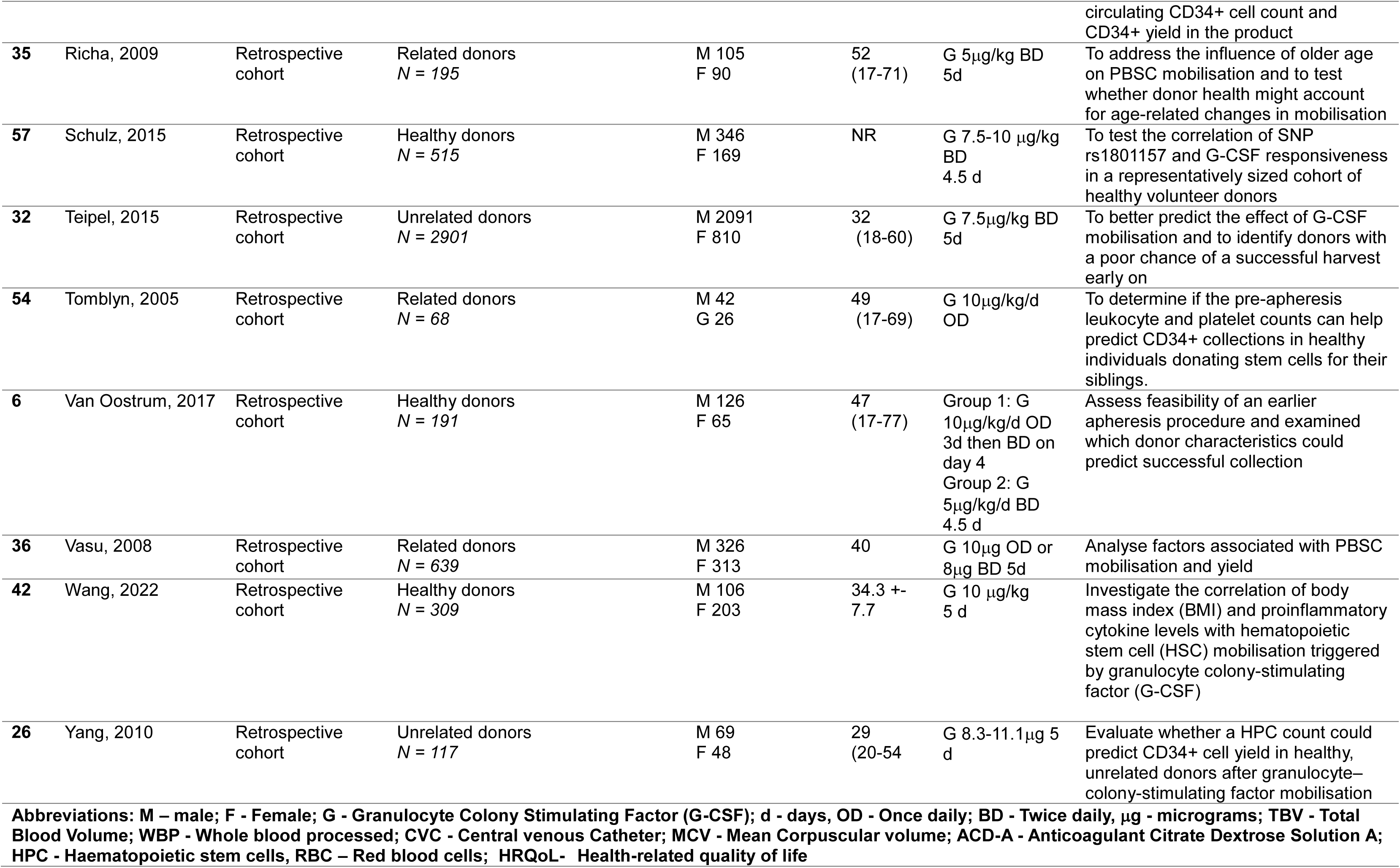
Characteristics of studies that assessed blood CD34+ cell concentration after G-CSF mobilisation.

**Table 2:** Characteristics of studies that assessed CD34+ cell yield after PBSC.

| **CD34<sup>+</sup> cell yield after G-CSF mobilisation studies** |  |  |  |  |  |  |  |  |
| --- | --- | --- | --- | --- | --- | --- | --- | --- |
| **Ref** | **Study** | **Study design** | **Population (unrelated donors, siblings)** | **Sex of donors** | **Age of donors Median (range)** | **Mobilisation regimen (dose, duration)** | **Apheresis collection procedure parameter** | **Study primary aim** |
| **Prospective and / or randomised study designs** |  |  |  |  |  |  |  |  |
| 11 | Basara, 2000 | Randomised split dose escalating study | Related donors<br>*N* = 36 | M 16<br>F 20 | 17-69 | Group 1 G 8µg/d split BD 4 or 5d<br>Group 2 G 11µg/d split BD 4 or 5d<br>Group 3 G 15µg/d split BD 4 or 5d | Fenwal CS-3000 Plus<br>3-4hr duration<br>3 x TBV | To determine whether escalating split doses of lenograstim result in increased number of CD34+ cells harvested |
| 29 | Beelan, 2002 | Prospective multicentre cohort | Related donors<br>*N* = 150 | M 75<br>F 75 | 42 (16-64) | G 10µg/kg/day OD OR<br>G 8µg/kg/day split into BD (depending on centre preference) | Continuous flow cell separator | Evaluate the feasibility of G-CSF mobilisation and collection of allogeneic PBSC in healthy related donors |
| 9 | Engelhardt, 2001 | Prospective cohort | Healthy donors<br>*N* = 52 | M 24<br>F 28 | 43 (20-61) | G 8µg/kg BD 4 d | Baxter CS 3000, or COBE Spectra, Intermediate LVA: 60-100ml/min<br>No CVC<br>TBV 2.5 (median 12L (3-18L))<br>Duration 2-3 hr | To determine the efficacy of intermediate dose G-CSF and intermediate large volume apheresis to mobilise PBSC for allogeneic transplant |
| 24 | Komeno, 2005 | Open labelled multicentre Randomised Controlled Trial | Related donors<br>*N* = 72 | M 40<br>F 31 | Grp 1: 43 (17-62)<br>Grp 2: 51 (19-65) | Group 1: G 400µg/m2 OD 4d<br>Group 2: 200µg/m2 BD 4d | COBE Spectra WBP 150-200mL/kg<br>Flow rate 60-80ml/min | Compare efficacy of once daily and twice-daily administration of filgrastim and evaluate the impact of potentially confounding factors on PBSC yield |
| 50 | Kroger, 2000 | Randomised control trial | Healthy donors<br>*N* = 50 | 29 M<br>21 F | Grp 1L Median 37 (21-64)<br>Grp2: Median 48 (27-56) | Group 1: 10µg/kg/d OR<br>Group 2: 5µg/kg/d BD | COBE Spectra 2.5 x TBV (8-14 L)<br>Flow rate 50-70ml/min | To confirm the schedule dependency of G-CSF administration in setting of PBSC mobilisation |
| 45 | Lysak, 2011b | Prospective cohort | Unrelated and related donors<br>*N* = 76 | M 53<br>F 23 | 39 (23-68) | G 10µg/kg OD 5 d | Not reported | To examine the changes in cytokine levels and adhesion molecule expression caused by |
|  |  |  |  |  |  |  |  | G-CSF, and correlation with CD34+ cell count |
| 49 | Martino, 2015b | Prospective cohort | Related donors<br>*N* = 75 | M 44<br>F 31 | 44.8+-<br>13.5 | G 10µg/kg OD compared to historical cohort of 181 HDs<br>5µg/kg BD 5 day | COM TEC cell separator in OD group, AS 104/AS 204 and COM.TEC in BD group<br>ACD-A | Compare the efficacy of peripheral blood HSPC Lenograstim once daily to twice daily |
| 43 | Pruszczyk, 2019 | Prospective cohort | Unrelated donors<br>*N* = 153 | M 100<br>F 53 | 28 (18-55) | G 5µg/kg BD 4 day then one dose am of apheresis | Spectra Optia | Evaluate the possible impact of repeated blood donation on efficiency of subsequent HPC mobilisation by granulocyte colony-stimulating factor (G-CSF) |
| **Retrospective study designs** |  |  |  |  |  |  |  |  |
| 10 | Al-Ali, 2011 | Retrospective cohort | Related donors<br>*N* = 167 | M 85<br>F 82 | 47<br>(18-74) | G 5µg/kg BD 5d | COBE Spectra | Analyse safety and the impact of age on mobilisation and collection of HPC |
| 8 | Almeida-Neto, 2019 | Retrospective Cohort | Unrelated donors<br>*N*= 338 | M 218<br>F 120 | 37<br>(IQR 28:46) | G 10µg/kg OD for 4-7 days (dose increased to max of 20µg/kg if inadequate mobilisation and some donors started with 5mcg/kg dose<br>G 10µg/kg 4-7 d OD | COBE Spectra 3-4x TBV<br>Blood flow 40-60ml/min<br>Duration 4-5hr | Validate a formula that predicted CD34+ cell yield and to describe variables that correlated with high yield mobilisation and collection in healthy donors |
| 51 | Anderlini, 2020 | Retrospective Cohort | Healthy donors<br>*N* = 81 | M 40<br>F 41 | Group A 43<br>Group B 44 | Group A: G 6µg/kg BD 3 d<br>Group B: 12µg/kg OD 3d | COBE Spectra 3 x TBV | Once daily Vs Twice daily G-CSF comparison |
| 22 | Basci, 2020 | Retrospective cohort | Healthy donors<br>*N* = 61 | M 38<br>F 23 | 54<br>(50-65) | 10µg/kg/day 4 or 5 d. If day 4 WCC>75 G stopped, if WCC >60 G halved | Continuous flow cell separator Fresenius Kabi 2-3 x TBV, ACD-A | To explore the effect of donors' age on stem cell harvesting, engraftment duration after Allo-SCT, and product quality. |
| 28 | Billen, 2014 | Retrospective cohort | Unrelated donors<br>*N* = 308 | M 247<br>F 61 | 32<br>(18-58) | G 10µg/kg 4d | COBE Spectra MNC technique 2-2.5 x TBV or Spectra Optia<br>Duration 4 hr<br>Median TBV 13.45L | To evaluate the frequency of unrelated PBSC collections that meet the target yield and the impact of donor factors on this. |
|  |  |  |  |  |  |  | ACD-A |  |
| 25 | Brissot, 2009 | Retrospective cohort | 69 related donors, 26 unrelated donors<br>*N* = 95 | M 50<br>F45 | 47 (18-81) | G 10µg/kg<br>5d | Not Reported | Analyse factors associated with the total CD34+ PBSC yield in an ethnically homogeneous european population |
| 46 | Chen, 2014 | Retrospective cohort | Related donors<br>*N* = 69 | M 40<br>F29 | 33 (19-59) | G 10µg/kg<br>5d | COBE Spectra<br>Minimum 12L TBV processed<br>ACD-A<br>Duration 4 hr | Further define donor factors that contribute to a good response to G-CSF mobilisation. |
| 12 | Farhadfar, 2020 | Retrospective cohort | Unrelated donors<br>*N* = 20884 | M 13365<br>F 7519 | 18-62 yrs | G 10µg/kg<br>5d | 12-24 L WBP | Examine the impact of BMI on PBSC mobilisation in a large cohort of adult unrelated donors |
| 47 | Farhadfar, 2022 | Retrospective cohort | Healthy donors<br>*N* = 336 | M 187<br>F 149 | 18-77 | G 11µg/kg<br>5d | 78% Large volume apheresis (>18L)<br>19% standard volume (12-<18L),<br>9% small (<12L) volume.<br>19% CV for access | Evaluate whether pre-donation HRQoL markers predict toxicity profile and stem cell yield following PBSC donation in healthy donors. |
| 7 | Flommersfield, 2015 | Retrospective cohort | Related donors<br>*N* = 23 | Day 5 group:<br>M 12<br>Day 4 group:<br>M 7 | Day5 Group:<br>Median 54 (27-74)<br>Day 4 Group:<br>Median 51 (28-67) | G 10µg/kg/d split into BD dosing 4 (n=18) or 5 d (n=25) | Spectra Optia | Investigate whether collection on Day 4 would lead to adequate amounts of PBPCs while minimizing granulocyte-colony-stimulating factor (G-CSF) exposure in healthy donors. |
| 44 | Hsu, 2015 | Retrospective cohort | Unrelated donors<br>*N* = 10776 | M 6778<br>F 3988 | 35 (18-61) | G 10µg/kg OD for 5 days (4 days before and am of collection) | Not Reported | Analyse the effect of donor race and ethnicity on PBSC mobilization in a large cohort of adult unrelated donors. |
| 13 | Janseen, 2012 | Retrospective cohort | Related donors<br>*N* = 223 | M 113<br>F 110 | 175< 61<br>61 > 61 (18-71) | G 10µg/kg<br>4d | COBE Spectra<br>MNC protocol<br>ACD-A and heparin<br>TBV decided by measuring number of CD34 cells collected after 1 blood volume had been processed. | Report experience of using older PBSC donors (61-71 years) and compare to younger donors (<61 years) who collected stem cells during the same time period. |
| 14 | Kayser, 2023 | Retrospective cohort | Healthy donors<br>*N* = 588 | M 463<br>F 125 | 29 (18-69) | G 10µg/kg<br>5d | Spectra Optia (Terumo) cMNC. | Identify factors predictive of successful CD34+ collection. In addition, we propose a refined |
|  |  |  |  |  |  |  | Duration 5h<br>maximum<br>ACD inlet ratio<br>14:1, inlet flow 40-<br>100ml/min | formula that predicted the minimal<br>CD34+ cell yield and took the<br>donor's and the recipient's body<br>weight into account. |
| 48 | Kroger, 2002 | Retrospective<br>cohort | Related and<br>unrelated donors<br>*N* = 90 | Group 1:<br>M 14<br>F 11<br>Group 2:<br>M 12<br>F 10 | Group 1:<br>37<br>Group 2:<br>48 | Group 1:5µg/kg BD<br>Group 2:8µg/kg BD | COBE Spectra<br>8-14 L WBP<br>Flow rate 50-70ml/min | Report experience of two dose<br>schedules of G-CSF and their<br>tolerability |
| 30 | Lenk, 2013 | Retrospective<br>Cohort | Unrelated donors<br>*N* = 463 | M 376<br>F 87 | 33<br>(18-54) | G 10µg/kg<br>5d | COBE Spectra<br>(97%)<br>Baxter CS 3000<br>(1%)<br>Fresenius Comtec<br>(23%)<br>ACD-A<br>4-5x TBV<br>50-110mL/min<br>3-4.5 hours | Investigate the assumed<br>association of the *CXCL12* gene<br>polymorphism and progenitor cell<br>mobilisation after G-CSF<br>administration |
| 15 | Lysak, 2005 | Retrospective<br>cohort | Healthy donors<br>(65 related, 46<br>unrelated)<br>*N* = 111 | M 68<br>F 43 | 40<br>(16-75) | G 10µg/kg<br>5-6 d | COBE Spectra<br>50-90ml/min<br>ACD-A<br>1.5-4.4 x TBV<br>processed (3.2<br>median)<br>Median Duration =<br>235 min | Evaluate the feasibility of G-CSF<br>mobilisation and collection of<br>allogeneic PBSC in healthy<br>donors |
| 16 | Lysak, 2011a | Retrospective<br>cohort | Unrelated and<br>related donors<br>*N* = 224 | M 144<br>F 80 | Older<br>group:<br>59 (55-75)<br>Younger<br>group:<br>29<br>(17-48) | G 11µg/kg/day OD or<br>BD 4 or 5 d | COBE Spectra<br>ACD-A,<br>CVC 42% older<br>donors and 26%<br>younger<br>2-3x TBV<br>Duration 4h | Report experience with mobilising<br>and collecting healthy donors by<br>using standardised procedures<br>and comparing donors aged ≥55<br>years with those <55 years old. |
| 23 | Martino, 2015a | Retrospective<br>cohort | Related donors<br>*N* = 246 | M 138<br>F 108 | Group1:<br>38<br>Group 2:<br>55<br>Group 3:<br>62<br>(18-70) | G 10µg/kg/d OD or<br>split dose BD 4 d | COM.TEC cell<br>seperators, AS<br>104/AS 204 and<br>COM.TEC in;<br>Fresenius Hemo-Care GmbH Bad<br>Homburg,<br>Germany. ACDA | Assess the efficacy and short and<br>long-term safety of G-CSF<br>mobilisation in an older healthy<br>donor population |
| 27 | Passeri, 2022 | Retrospective cohort | Healthy donors<br>*N* = 804 | M 447<br>F 393 | 45<br>(16-74) | G 10µg/kg split dose 4 or 5 d | COBE Spectra (Terumo), CS-3000 Blood cell separator, COM.TEC or Amicus<br>Number of cycles and PBV adjusted to achieve target CD34+ cell dose of 4x10<sup>6</sup>/kg recipient body weight | Investigate whether haematopoietic progenitor cell apheresis on the 4th day after the first dose of G-CSF might lead to an adequate harvesting of stem cells, reducing G-CSF exposure. |
| 52 | Pinar, 2022 | Retrospective cohort | Unrelated donors<br>*N* = 142 | M 102<br>F 40 | 33 (19-52 range) | G 5µg/kg 12 BD (n = 83) Vs 10µg/kg/OD (n = 59).<br>Neupogen (n = 106) and Tevagrastim (n = 36) | Spectra Optia (Terumo BCT). TBV 100-150ml/kg 40-50mL/min flow rate, | Compare the efficacy of both once-daily and divided-dose G-CSF administration and the original and biosimilar GCSF use and the frequency and severity of adverse events in donors. |
| 21 | Pornprasertsud, 2015 | Retrospective cohort | Related donors<br>*N* = 50 | M 25<br>F 25 | 38.5 (17-65) | G 10µg/kg 5d | COBE Spectra 40-60mls/min<br>Duration 5 hr | Analyse the predicting factors for high number of peripheral blood CD34+ cells |
| 56 | Pruszczyk, 2020 | Retrospective cohort | Unrelated donors<br>*N* = 330 | M 214<br>F 116 | 29 (18-56) | G | Spectra optia (Terumo)<br>ACD-A | To assess the possible impact of seasonal variation in the response to G-CSF in healthy hematopoietic stem cell donors |
| 17 | Ramaligham, 2021 | Retrospective cohort | Healthy donors<br>*N* = 100 | M 70<br>F 30 |  | G 10µg/kg 5d | Spectra Optia CMNC<br>ACD-A<br>2.5-3 x TBV processed (10-12L)<br>Flow rate 150-180 min | Evaluate the impact of pre apheresis haematological variables on CD34+ yield. |
| 18 | Rinaldi, 2012 | Retrospective cohort | 174 related donors, 16 unrelated donors<br>*N* = 190 | M 105<br>F 85 | 44 (18-72) | G 10µg/kg/d 4-6 d<br>If haploidentical donor 12-16µg/kg/d 4-5 d | COBE Spectra MNC protocol 2x TBV | The Evaluate the safety of PBSC donation in related and unrelated donors during short- and long-term follow-up and to analyse factors affecting CD34+ mobilisation capacity and collection efficacy, to optimise donor selection. |
| 6 | Van Oostrum, 2017 | Retrospective cohort | Healthy donors<br>*N* = 191 | M 126<br>F 65 | 47 (17-77) | Group 1: G 10µg/kg/d OD 3d then BD on day 4 | Spectria Optia 14-15.3L processed blood volumes | Assess feasibility of an earlier apheresis procedure and examined which donor |

|  |  |  |  |  |  | Group 2: G 5µg/kg/d<br>BD 4.5 d | 31-58 mL/min flow rate<br>Max duration 6 h | characteristics could predict successful collection |
| --- | --- | --- | --- | --- | --- | --- | --- | --- |
| **19** | Wang, 2008 | Retrospective cohort | Unrelated donors<br>*N* = 375 | M 229<br>F 146 | 31.2<br>(20-54) | G 10µg/kg<br>5d | COBE Spectra<br>40-60ml/min | Gain insight into the donor factors that may affect quantity of stem cell collection from volunteer donors. |
| **55** | Wang, 2013 | Retrospective cohort | Healthy donors<br>*N* = 793 | M 494<br>F 302 | 33 in low MCV group<br>31 in normal MCV group | G 10µg/kg<br>5d | COBE Spectra<br>40-60ml/min<br>Target processing volume based on recipient weight:<br>12L for <35kg, 15L for 35-45kg and 18L if >45kg | Evaluate the impact of low MCV of RBCs on the outcome of stem cell mobilisation and leukapheresis |
| **26** | Yang, 2010 | Retrospective cohort | Unrelated donors<br>*N* = 117 | M 69<br>F 48 | 29<br>(20-54) | G 8.3-11.1µg<br>5 d | COBE spectra<br>Large volume apheresis<br>40-60mL/min<br>6-8 hours duration | Evaluate whether a HPC count could predict CD34+ cell yield in healthy, unrelated donors after granulocyte–colony-stimulating factor mobilisation |
| **20** | Zhen, 2019 | Retrospective cohort | Unrelated donors<br>*N* = 145 | M 145<br>F 0 | 37 | G 5-10µg/kg BD<br>5d | COBE Spectra<br>TBV x 3<br>49.5-70mk/min | To investigate the correlation between smoking load and PBSCs mobilisation by G-CSF in healthy donors. |

**Abbreviations:** M – male; F - Female; G - Granulocyte Colony Stimulating Factor (G-CSF); d - days, OD - Once daily; BD - Twice daily, µg - micrograms; TBV - Total Blood Volume; WBP - Whole blood processed; CVC - Central venous Catheter; MCV - Mean Corpuscular volume; ACD-A - Anticoagulant Citrate Dextrose Solution A; HPC - Haematopoietic stem cells, RBC – Red blood cells; HRQoL- Health-related quality of life

### Study characteristics

Forty-one (80%) of the 51 eligible studies were retrospective cohort design. Seven (14%) of the remaining studies used prospective data collection but only three (6%) were prospective randomised trials that evaluated the effects of alternative G-CSF dose and/or scheduling. The eligible studies included at total of 44,759 doors but individual studies ranged in size between 23 to 20,884 donors. Eight studies included >500 donors and 14 included <100 donors. The median donor ages ranged from 29 to 59 years. 72% of all donors were male.

#### 1. Donor factors that influence blood CD34+ concentration or yield

##### Age

Donor age was the most frequently assessed factor in the eligible studies. Of the 25 studies that reported the relationship between age and CD34^+^ cell yield 16 (64%) reported an association between age > 40 years and lower CD34^+^ cell yield (6–21). Four further studies identified a non-significant trend towards this association (22–25) whereas five studies reported no association (26–30). Of the 25 studies that assessed the relationship between age and blood CD34^+^ cell concentration, 13 (52%) reported an association between older age and lower blood CD34^+^ cell concentration (8–10, 13, 16, 17, 21, 31–36). Eight further studies reported a similar trend but without statistical significance (6, 22, 23, 37–41) whereas in three studies, there was no association (26, 27, 42). The final study reported that older age was associated with higher blood CD34^+^ cell concentration (30). Analysis of the significance of donor age was hampered by the variation between the studies in whether age was considered as a continuous or categorical factor and in the categorical definitions of ‘older age’ (range >30 to >60 years). The median and range of included donors varied considerably between studies (Table 1 and 2).

##### Gender

Of the 17 studies that assessed the association between CD34^+^ cell yield and donor gender 12 (71%) reported higher CD34+ cell yields in males (9, 10, 12, 14, 17–19, 25, 28, 30, 43, 44) but the remaining five studies (29%) found no association (6, 15, 16, 24, 29). Of the 20 studies that assessed blood CD34+ cell concentration, eight (42%) reported higher concentrations in males (9, 30, 32, 34, 36, 37, 39, 45) whereas 12 (59%) reported no significant effect of gender (6, 17, 21, 22, 31, 33, 35, 38, 40–42, 46).

##### Ethnicity

Only four studies assessed associations between donor ethnicity and CD34+ cell yield from PBSC or blood CD34+ cell concentration. Three of these reported that compared to European ancestry donors, African ancestry (AA) donors had higher CD34+ cell yields (12, 44) and blood CD34+ cell concentrations (35, 36, 44). In one of these studies that included >10,000 donors, the effect of AA was most pronounced in male donors with higher body mass index (BMI) (44). The remaining study reported a trend of a higher blood CD34+ cell concentrations in AA donors but contained only 16 subjects with this ancestry (35).

##### Anthropometric characteristics

Of the nine studies that assessed the association between BMI and CD34+ cell yield, four reported that high BMI was associated with higher yield (6, 12, 19, 46). Seven of nine studies reported an association between high BMI and higher blood CD34+ concentration (6, 26, 30, 32, 37, 42, 46). None of the seven studies that assessed donor height reported an association with either CD34+ cell yield (20, 25, 29), or blood CD34+ cell concentration (36, 38, 40, 41). All four of the studies that assessed the differences in weight between donor and recipients reported an association between donors having greater positive weight differences and higher CD34^+^ cell yield (6, 8, 14, 28). Higher donor blood volume estimated using the Nader method was associated with higher blood CD34+ cell concentration in one study (41), but not in another (40).

##### Other donor factors

One of two studies that assessed the effect of donor smoking reported an association with lower CD34+ cell yield (20), whereas the other found no effect (43). Two studies found no association between smoking and CD34+ cell concentration (30, 39) but in one retrospective study, there was an association between smoking and lower concentration (32). Single studies reported no associations between donor comorbidity expressed as the Charlson Comorbidity Index and CD34^+^ cell concentration (35) and between previous blood donation or physical activity and CD34^+^ cell yield (43). One study reported no association between self-reported health-related quality of life and CD34^+^ cell yield (47).

#### 2. Mobilisation factors that influence blood CD34+ concentration or yield

##### G-CSF dose

Seven studies assessed the impact of G-CSF dose on CD34^+^ cell yield. In an open label randomised split dose escalating study in 36 donors, CD34+ cell yield was the same between three G-CSF dose groups (8, 11 or 15 μg/kg/day split into two doses), although the 15μg group had PBSC on day 4 of G-CSF treatment whilst the 8μg and 11μg groups had PBSC on day 5 (11). In a second trial, 150 donors were randomised to receive either 10μg/kg/day G-CSF once daily or 8μg/kg/day split into two doses. The donors receiving 10μg/kg/day once daily were more likely to achieve a target CD34+ cell yield of >5×10^6^/kg recipient weight. The frequency of adverse events was similar between groups (29). Of the five remaining studies that were retrospective, one reported that treatment with 8μg/kg twice per day G-CSF resulted in higher CD34+ cell yields compared to 5μg/kg twice per day but was associated with more frequent bone pain, headache and fatigue (48). In a second study, all donors received G-CSF doses of 10μg/kg/day administered either as a single dose or split between two doses. Amongst the donors who achieved a satisfactory CD34+ cell yield (>5×10^6^ cells/Kg), a higher total G-CSF dose was reported in donors receiving twice daily G-CSF but not once daily G-CSF (6). In the three remaining retrospective studies, all the donors were prescribed the same 10μg/kg/day yet received different actual doses because of nearest whole vial dosing. None of these studies showed a relationship between actual G-CSF dose received and CD34+ cell yield (16, 19, 44).

Eight studies assessed the impact of G-CSF dose on blood CD34^+^ concentration, all of which were retrospective. In one study, 16μg/kg/day G-CSF resulted in higher blood CD34+ cell concentrations than 10μg/kg/day (36). In two studies, higher blood CD34+ concentrations were associated with higher total G-CSF doses received by donors prescribed 10μg/kg/day G-CSF rounded to the nearest whole vial size (6, 38).The remaining five studies found no association between blood CD34+ cell concentration and higher G-CSF doses (16, 31, 34, 37, 39). One of these studies found an increased frequency of bone pain, headaches, and chills with G-CSF doses >10μg/kg/day (37).

##### G-CSF scheduling

The effect once daily (OD) vs twice daily (BD) G-CSF dosing on CD34^+^ cell yield was assessed in five studies. In one prospective study in 72 donors, G-CSF 400μg/m^2^ OD and 200μg/m^2^ BD resulted in similar CD34+ cell yields (24). Comparison of CD34+ cell yields in 75 donors receiving G-CSF 10μg/kg OD to a historical control group who received G-CSF 5μg/kg BD also found no difference between the groups (49). By contrast, in 50 donors receiving either 10μg/kg OD or 5μg/kg BD, there was a higher CD34+ cell yield in the BD group (50). The remaining two retrospective studies found no differences in CD34+ cell yields in donors who received OD or BD G-CSF scheduling (51, 52). There were no reported associations between G-CSF scheduling and blood CD34^+^ cell concentration in two retrospective studies (16, 52) and one prospective study (49).

The impact of PBSC on day four compared to day five of G-CSF treatment on CD34+ cell yield was assessed in three retrospective studies. In one study, day 4 PBSC, resulted in insufficient CD34+ cell yield in most donors whereas this was not the case for day 5 PBSC (6). In contrast, a second study reported no difference between day 4 and day 5 PBSC (7). In the final retrospective study of 840 donors, the CD34+ cell yield was lower in donors undergoing day 4 PBSC compared to day 5 PBSC, although both groups were equally likely to reach a target CD34+ cell yield of >4×10^6^/kg recipient weight (27). This same study also reported that CD34^+^ cell concentration was higher at the day 5 PBSC compared to day 4 (27). In one retrospective study, lymphapheresis prior to G-CSF mobilisation increased CD34^+^ cell concentration after five days of G-CSF mobilisation (36).

#### 3. Collection factors that influence blood CD34+ concentration and yield

Amongst the eligible studies that reported collection factors, there was wide variation in collection process. The most frequently used apheresis machine was the COBE spectra. Although the total reported processed blood volume (PBV) was typically 2.5 to 3 total blood volume some studies reported up to 5 blood volumes in a single PBSC (30). ACD-A was the anticoagulant used in all studies which reported this data.

The use of a central line for PBSC venous access was reported to increase CD34+ cell yield in one retrospective study (19) but was found to have no effect in a second study except in a sub-group of female donors where a central line increased yield compared to peripheral venous access (44). The PBV was assessed in six studies, of which two were prospective (24, 29). In one of these, a PBV of >200mls/kg was associated with higher CD34+ cell yield compared with lower PBVs. In contrast, the second prospective study of donors who had a PBV in the range 150-200mls/kg, reported there was no relationship between the exact PBV and CD34+ cell yield. Three of the remaining four retrospective studies, reported that a higher PBV was associated with higher CD34^+^ cell yield (20, 28, 46). In contrast, the remaining retrospective study of 20,884 donors found that higher PBV was associated with a lower CD34^+^ yield (12). This was attributed to declining collection efficacy with larger collection volumes. In one retrospective study, faster flow rate during PBSC was associated with a lower CD34^+^ cell yield (19).

#### 4. Laboratory factors

Amongst the eligible studies, the most commonly reported laboratory factors were full blood count (FBC) parameters and blood CD34+ cell concentration in blood samples collected before the start of G-CSF mobilisation (baseline) or after mobilisation but before PBSC (pre-PBSC). In all 16 studies that assessed the pre-PBSC blood CD34+ concentration, there was a consistent association with the subsequent CD34^+^ cell yield from the first PBSC collection (7–9, 11, 14, 15, 18–21, 24–26, 45, 46).

##### Pre-PBSC full blood count parameters

The association between pre-PBSC white cell count (WCC) and CD34^+^ cell yield was inconsistent amongst the eight studies that evaluated this parameter (7–10,12,14,16,41) with only three reporting a significant positive association (12,14,41). Five studies found that higher WBC was associated with higher CD34+ cell concentrations in the same pre-PBSC blood sample (8, 31, 34, 53, 54), whereas two reported no association (17, 33). Higher pre-PBSC platelet (PLT) count was associated with higher CD34+ cell yield in four out of ten studies that assessed yield (12, 15, 16, 21). In four studies, higher PLT was associated with higher CD34+ cell concentrations in the same pre-PBSC blood sample (8, 31, 33, 54), whereas there was no association in one further study (17). Higher pre-PBSC haemoglobin concentration (HB) was associated with higher CD34+ cell yield in two of the four studies that evaluated yield (8, 25). Pre-PBSC HB was not associated with CD34+ cell concentration in two studies (17, 33).

Higher pre-PBSC haematocrit (HCT) (17, 21), neutrophil (NEUT) count (12, 17) and nucleated red cells (nRBC) counts (17) were also associated with increased CD34+ cell yield, but not with higher CD34+ cell concentration in the pre-PBSC blood sample (17).

##### Baseline full blood count parameters

Fewer studies have assessed baseline FBC parameters. Of four studies that assessed CD34+ cell yield, two studies reported an association between higher baseline WCC and higher yield (8, 24), including a randomised control trial in which baseline WCC was the strongest predictor of being a ‘poor mobiliser’ (defined as CD34+ cell yield < 2×10^6^/kg donor weight). Two studies found no association (20, 25). Higher baseline WCC was associated with higher CD34+ cell concentration in one out of five studies that assessed concentration (31) whereas there was no association in the other studies MARTINO 2006, (32, 34, 40, 41).

Higher baseline NEUT count and CD34+ cell count were associated with higher CD34+ cell yield in two studies (12, 18). Higher baseline CD34+ count was associated with higher CD34+ cell concentration in the pre-PBSC blood sample (40, 41). Higher baseline PLT count was associated with higher CD34+ cell yield in one retrospective study (12), but not in another (25). Higher basline PLT count was also associated with higher CD34+ concentration in the pre-PBSC sample in three retrospective and one prospective study (32, 36, 39, 40), but not in other studies (17, 41).

Higher baseline HB (8, 25), and mean red cell volume (MCV) (55) were associated with higher CD34+ cell yield in two retrospective studies. Baseline HB was also associated with CD34+ cell concentration in the pre-PBSC sample in one retrospective study (39) but was there was no association in three other studies (32, 41, 49). Higher baseline MCV was not associated with CD34+ concentration in the pre-PBSC sample in two retrospective studies (21, 39). Amongst a range of other FBC parameters evaluated in a retrospective study of 333 donors, only higher baseline mononuclear count was associated with CD34+ cell concentration in the pre-PBSC sample (36).

##### Other factors

Possible associations between CD34+ cell concentration in the pre-PBSC sample and baseline tests for clotting, liver, and renal function, and with baseline CRP, cholinesterase (CHE), lactate dehydrogenase (LDH), ferritin and immunoglobulin concentrations were assessed by a single retrospective study. Only higher CHE and higher LDH were significantly associated with higher CD34+ cell concentration in the pre-PBSC sample (39) Neither other cell surface markers on CD34+ cells nor baseline inflammatory cytokine concentrations were associated with CD34+ cell yield in one study (14) whereas in a different retrospective study higher CD11a expression on CD34+ cells in the collected cells was associated with lower CD34+ concentration in the pre-PBSC sample (45). There was a non-significant trend towards lower CD44 expression on CD34+ cells from donors with a lower CD34+ concentrations (45). In two studies that investigated the relationship between baseline inflammatory cytokine concentrations and CD34+ concentration in the pre-PBSC sample (42, 45), both reported that higher TNFα concentration was associated with higher CD34+ cell concentration. Higher IFNγ and IL-22 were also associated in one study (42) but not in another (45).

The seasonal timing of mobilisation and cell collection was assessed in a single retrospective study but was not associated with CD34+ cell yield (56).The effects of the a common haplotype in *CXCL12* on both CD34+ concentration and yield were assessed in a single retrospective study of 463 healthy donors but no significant associations were identified (30, 57).

## DISCUSSION

This scoping review offers a comprehensive overview of the donor, mobilisation, collection and laboratory factors that are associated with favourable donation outcomes from standard G-CSF mobilisation and PBSC in allogeneic stem cell donors. The review identifies factors associated with CD34+ cell concentration in donor blood immediately before PRBC and also the yield of CD34+ cells from the subsequent collection procedure. These alternative endpoints are closely related as multiple previous studies found that blood CD34+ cell concentration before PBSC was consistently associated with the subsequent yield. One striking observation from this analysis was that previously performed association studies are heterogeneous and frequently are constrained by small sample size. Despite this, there are sufficient data to identify several important association trends with these donation outcomes.

The long-known association between younger donor age and higher stem cell yield (29), was confirmed in multiple studies assessed in this review irrespective of whether the outcome was higher CD34+ cell concentration or yield. This finding aligns with the conclusion of a recent review of clinical and mobilisation factors (29) although some uncertainty remains because of the inconsistent definitions of ‘older age’ in prior studies and whether age was considered as a continuous or categorical variable. In the context of expanding demand for allogeneic stem cell donations, there is an increasing requirement to consider older donors. Although evidence from most previous studies raises concern about reduced yield, some studies showed that despite reduced absolute yields from older donors, this did not commonly result in inadequate or failed donations (10, 22, 25). However, safety concerns about using older donors have been raised elsewhere, particularly concerning diminishing telomere length, clonal haematopoiesis risk, and regenerative potential decline in stem cell from older donors (58, 59). CD34+ yield alone may not be the only relevant marker of successful stem cell collection from older donors, suggesting a need for more sophisticated selection algorithms.

In the studies that assessed donor gender, being male was generally associated with higher stem cell yield possibly because of higher donor weight and therefore G-CSF dosing in male compared to female donors (36). Our review also identified that being male was more frequently associated with higher CD34+ cell yield than with higher CD34+ concentration in blood before PBSC. This suggests that part of the effect of being male is at stem cell collection, possibly because male donors typically having higher circulating blood volume enabling higher processed blood volumes during PBSC which was associated with high yield in multiple studies (20, 24, 28, 29, 46). Donor African ancestry was associated with higher stem cell yield in some, but not all studies. Associations with ethnicity were sometimes inferred indirectly from studies not primarily designed to examine the effect of this donor factor (35), raising the possibility of bystander effect or bias. The strongest evidence of an effect from ethnicity comes from a retrospective study of 10,776 donors which demonstrated higher CD34+ cell yields in African compared to European ancestry donors, particularly in donors with high BMI (44). This effect was not explained by the higher G-CSF doses received by high BMI donors (44).

Laboratory factors were assessed less frequently than donor clinical factors in the eligible studies included in this review. Our finding of a consistent association between CD34+ concentration in blood before PBSC and subsequent CD34+ cell yield is an important finding as it confirms the importance of measuring CD34+ cell concentration after mobilisation to inform decision making about whether to proceed with PBSC or whether to extend or escalate mobilisation. The two studies identified that identified an association between baseline blood CD34+ cell concentration and CD34+ cell concentration after mobilisation (40, 41) are of particular interest since these suggest that laboratory prediction of stem cell yield could even occur before the start of mobilisation potentially enabling greater personalisation of mobilisation protocols by the addition of other agents such as Plerixafor (60).

Other laboratory factors often associated with CD34+ cell concentration or yield included baseline or pre-PBSC WCC, PLT count and HB concentration. In contrast to analysis of blood CD34+ cell concentration, these are widely and rapidly accessible as part of the FBC test. Mechanistically, these associations may reflect commonality between cellular pathways that mediate CD34+ cell mobilisation and those which regulate the maturation of stem cell into mature blood cells, as has been suggested previously for blood PLT (36). Although our review has identified promising signals for the use of pre-PBSC or even baseline FBC parameters in predicting favourable donation outcomes, the most consistent evidence was for associations with blood CD34+ cell concentration rather than yield after PBSC. This apparent discrepancy could be explained by the comparatively small number of studies examining associations with CD34+ cell yield, possibly hampering identification of significant associations with this outcome. Further evidence from larger, prospective study particularly of the CD34+ cell yield outcome is required to confirm clinical utility.

Mobilisation and collection factors associated with higher stem cell yields are attractive to identify since these are readily implementable into modified donor protocols. However, amongst the studies evaluated in this review, there were inconsistent findings about the impact of G-CSF dose or dose scheduling on yield but some evidence of an adverse safety signal with higher G-GSF doses (29). G-CSF has a rapid bioavailability resulting in peak plasma levels four hours after administration (61) and a short plasma half-life. Although this suggests a potential advantage of twice daily G-CSF dosing, the pharmacodynamic effect of G-CSF on circulating cell counts is frequently longer than 48 hours, suggesting that twice daily dosing may not be necessary. Moreover, twice daily G-CSF dosing is less tolerable to donors because of injection frequency. Unfortunately, we cannot confirm from existing literature whether once or twice daily dosing is superior for stem cell yield. There was greater consensus indicating superiority of five rather than four days of G-CSF treatment which is in keeping with longitudinal measurement studies of blood CD34+ cell concentrations which suggest that peak cell concentrations are reached by day 5 (27). Despite multiple previous studies, it is difficult to reach robust conclusions about the relationship between collection factors and CD34+ cell yield.

Potentially informative donor factors associated with donation outcomes in adults that are conspicuously absent from the literature are genetic factors, with only a single study examining a potential association with the *CXCL12* gene (30). This is surprising since there is abundant evidence elsewhere of a significant genetic component to CD34+ cell yield. This includes the murine models with strain specific variation in CD34+ yield after mobilisation associated with loci on chromosomes 2 and 11 (62). Several human candidate gene association studies have also been performed in paediatric or autologous donor populations that implicate multiple loci implicated biologically in the mechanism of G-CSF-induced stem cell mobilisation. These include variants in *VCAM1* (62), *CSF3R* (63, 64), *VLA4* (65), *CD44* (66), and *CXCL12* (57, 67). The importance of variation at these and other potential loci for stem cell mobilisation in adult allogeneic donors remains uncertain but is likely to be an area of future research study.

The strengths of this scoping review are its comprehensiveness and that we have considered two different outcomes of stem cell collection. These include CD34+ cell concentration before PBSC which could potentially be influenced by donor and mobilisation factors. The other outcome, CD34+ cell yield may be influenced by these factors, but also by collection factors and is also the outcome that is directly relevant for recipient outcomes. The main limitation of this review is that even though we have used a scoping strategy, inferences about many candidate factors remain uncertain because of the abundance of retrospective studies with small sample sizes, and the variable definitions of candidate factors and outcomes. A separate review has recently been published that also examined factors associated with stem cell yield, although this only examined associations with CD34+ concentration before PBSC (31). The conclusions of this review largely accord with our study although since we have also examined CD34+ cell yield, we have been able to resolve additional associations with collection factors arising from the process of PBSC.

Our findings are important because they highlight the need for future research particularly focussing on factors such as donor ethnicity and the genetic predictors of yield that are incompletely resolved at present. The future discovery of genetic predictors of successful donation is particularly attractive since it potentially enables better prediction of likely good or poor donors using genetic data at the point donors join registries rather than after donors have undergone mobilisation or collection. Future prospective data collection will enable better identification of predictors of successful donation and better use of these data through approaches such as machine learning (68).

## CONCLUSION

Younger age, male gender and having a blood higher CD34+ concentration immediately before PBSC are confirmed as the most robust predictors of CD34+ cell yield following G-CSF mobilisation and PBSC. Many other clinical, mobilisation, collection and laboratory factors show some evidence of association with higher CD34+ cell yield yet require stronger evidence from large, prospective studies to confirm their clinical utility.

## AUTHOR CONTRIBUTIONS

SS, JG, KS and RP contributed to the design of the study. CD contributed to search strategy design. KB and RP contributed to data curation. RP and AKW contributed to data curation. RP, AM, KS contributed to writing the manuscript.

## Data Availability

All data produced in the present work are contained in the manuscript

## REFERENCES

1. Holig K. G-CSF in Healthy Allogeneic Stem Cell Donors. Transfus Med Hemother. 2013;40(4):225–35.

2. Holig K, Kramer M, Kroschinsky F, Bornhauser M, Mengling T, Schmidt AH, et al. Safety and efficacy of hematopoietic stem cell collection from mobilized peripheral blood in unrelated volunteers: 12 years of single-center experience in 3928 donors. Blood. 2009;114(18):3757–63.

3. Pulsipher MA, Chitphakdithai P, Logan BR, Leitman SF, Anderlini P, Klein JP, et al. Donor, recipient, and transplant characteristics as risk factors after unrelated donor PBSC transplantation: beneficial effects of higher CD34+ cell dose. Blood. 2009;114(13):2606–16.

4. Remberger M, Gronvold B, Ali M, Mattsson J, Egeland T, Lundin KU, et al. The CD34(+) Cell Dose Matters in Hematopoietic Stem Cell Transplantation with Peripheral Blood Stem Cells from Sibling Donors. Clin Hematol Int. 2020;2(2):74–81.

5. Tricco AC, Lillie E, Zarin W, O’Brien KK, Colquhoun H, Levac D, et al. PRISMA Extension for Scoping Reviews (PRISMA-ScR): Checklist and Explanation. Ann Intern Med. 2018;169(7):467–73.

6. van Oostrum A, Zwaginga JJ, Croockewit S, Overdevest J, Fechter M, Ruiterkamp B, et al. Predictors for successful PBSC collection on the fourth day of G-CSF-induced mobilization in allogeneic stem cell donors. Journal of Clinical Apheresis. 2017;32(6):397–404.

7. Flommersfeld S, Sohlbach K, Jaques G, Bein G, Hoffmann J, Kostrewa P, et al. Collection of peripheral blood progenitor cells on Day 4 is feasible and effective while reducing granulocyte-colony-stimulating factor exposure to healthy donors. Transfusion. 2015;55(6):1269–74.

8. Almeida-Neto C, Rocha V, Moreira FR, Hamasaki DT, Farias MC, Arrifano AM, et al. Validation of a formula predictive of peripheral blood stem cell yield and successful collection in healthy allogeneic donors. Hematol Transfus Cell Ther. 2020;42(2):164–5 e5.

9. Engelhardt M, Bertz H, Wasch R, Finke J. Analysis of stem cell apheresis products using intermediate-dose filgrastim plus large volume apheresis for allogeneic transplantation. Annals of Hematology. 2001;80(4):201–8.

10. Al-Ali HK, Bourgeois M, Krahl R, Edel E, Leiblein S, Poenisch W, et al. The impact of the age of HLA-identical siblings on mobilization and collection of PBSCs for allogeneic hematopoietic cell transplantation. Bone Marrow Transplantation. 2011;46(10):1296–302.

11. Basara N, Schmetzer B, Blau IW, Bischoff M, Gunzelmann S, Kirsten D, et al. Lenograstim-mobilized peripheral blood progenitor cells in volunteer donors: an open label randomized split dose escalating study. Bone Marrow Transplantation. 2000;25(4):371–6.

12. Farhadfar N, Hsu JW, Logan BR, Sees JA, Chitphakdithai P, Sugrue MW, et al. Weighty choices: selecting optimal G-CSF doses for stem cell mobilization to optimize yield. Blood advances. 2020;4(4):706–16.

13. Janssen WE, Rahn D, Hackett M, Coyle D, Tomblyn M, Smilee RC, et al. Apheresis and transplant of hematopoietic progenitor cells (HPC) from allogeneic donors of age above 60 years. Bone Marrow Transplantation. 2012;47(12):1520–5.

14. Kayser S, Schlenk RF, Steiner M, Kluter H, Wuchter P. Predicting Successful Hematopoietic Stem Cell Collection in Healthy Allogeneic Donors. Transfus Med Hemother. 2023;50(5):396–402.

15. Lysak D, Koza V, Jindra P. Factors affecting PBSC mobilization and collection in healthy donors. Transfusion & Apheresis Science. 2005;33(3):275–83.

16. Lysak D, Koristek Z, Gasova Z, Skoumalova I, Jindra P. Efficacy and safety of peripheral blood stem cell collection in elderly donors; does age interfere? Journal of Clinical Apheresis. 2011;26(1):9–16.

17. Ramalingam TR, Vaidhyanathan L, Muthu A, Prabhakar V, Ramakrishnan B, Raj R, et al. Simple predictors of peripheral blood stem cell yield in healthy donors: A retrospective analysis in a tertiary care hospital. Journal of Applied Hematology. 2021;12(1):31–6.

18. Rinaldi C, Savignano C, Pasca S, Sperotto A, Patriarca F, Isola M, et al. Efficacy and safety of peripheral blood stem cell mobilization and collection: a single-center experience in 190 allogeneic donors. Transfusion. 2012;52(11):2387–94.

19. Wang TF, Wen SH, Chen RL, Lu CJ, Zheng YJ, Yang SH, et al. Factors associated with peripheral blood stem cell yield in volunteer donors mobilized with granulocyte colony-stimulating factors: the impact of donor characteristics and procedural settings. Biology of Blood & Marrow Transplantation. 2008;14(11):1305–11.

20. Zhen C, Fang X, Ding M, Wang X, Yuan D, Sui X, et al. Smoking is an important factor that affects peripheral blood progenitor cells yield in healthy male donors. Journal of clinical apheresis. 2020;35(1):33–40.

21. Pornprasertsud N, Niparuck P, Kidkarn R, Puavilai T, Sirachainan N, Pakakasama S, et al. The use of hematocrit level for predicting the efficiency of peripheral blood CD34(+) cell collection after G-CSF Mobilization in Healthy Donors. J Clin Apher. 2015;30(6):329–34.

22. Basci S, Bakirtas M, Uncu Ulu B, Yigenoglu TN, Yaman S, Batgi H, et al. Old is bad? The effect of age on peripheral stem cell mobilization and transplantation outcomes. Transfusion and apheresis science : official journal of the World Apheresis Association : official journal of the European Society for Haemapheresis. 2020:103007-.

23. Martino M, Bonizzoni E, Moscato T, Recchia AG, Fedele R, Gallo GA, et al. Mobilization of hematopoietic stem cells with lenograstim in healthy donors: efficacy and safety analysis according to donor age. Biology of Blood & Marrow Transplantation. 2015;21(5):881–8.

24. Komeno Y, a Y, Hamaki T, Mitani K, Iijima K, Ueyama J, et al. A randomized controlled trial to compare once-versus twice-daily filgrastim for mobilization of peripheral blood stem cells from healthy donors. Biology of Blood & Marrow Transplantation. 2006;12(4):408–13.

25. Brissot E, Chevallier P, Guillaume T, Delaunay J, Ayari S, Dubruille V, et al. Factors predicting allogeneic PBSCs yield after G-CSF treatment in healthy donors. Bone Marrow Transplantation. 2009;44(9):613–5.

26. Yang SH, Wang TF, Tsai HH, Lin TY, Wen SH, Chen SH. Preharvest hematopoietic progenitor cell counts predict CD34+ cell yields in granulocyte-colony-stimulating factor-mobilized peripheral blood stem cell harvest in healthy donors. Transfusion. 2010;50(5):1088–95.

27. Passeri C, Iuliani O, Di Ianni M, Sorrentino C, Giancola R, Abbruzzese L, et al. Comparison between peripheral blood progenitor cell collection on the 4(th) or 5(th) day of granulocyte colony-stimulating factor treatment in allogeneic stem cell donors: implications for hematopoietic progenitor cell apheresis guidelines. Blood Transfus. 2023;21(1):37–41.

28. Billen A, Madrigal JA, Szydlo RM, Shaw BE. Female donors and donors who are lighter than their recipient are less likely to meet the CD34+ cell dose requested for peripheral blood stem cell transplantation. Transfusion. 2014;54(11):2953–60.

29. Beelen DW, Ottinger H, Kolbe K, Ponisch W, Sayer HG, Knauf W, et al. Filgrastim mobilization and collection of allogeneic blood progenitor cells from adult family donors: first interim report of a prospective German multicenter study. Annals of Hematology. 2002;81(12):701–9.

30. Lenk J, Bornhauser M, Kramer M, Holig K, Poppe-Thiede K, Schmidt H, et al. Sex and body mass index but not CXCL12 801 G/A polymorphism determine the efficacy of hematopoietic cell mobilization: a study in healthy volunteer donors. Biol Blood Marrow Transplant. 2013;19(10):1517–21.

31. Piccirillo N, Putzulu R, Metafuni E, Massini G, Fatone F, Corbingi A, et al. Peripheral Blood Allogeneic Stem Cell Mobilization: Can We Predict a Suboptimal Mobilization? Transfus Med Rev. 2023;37(2):150725.

32. Teipel R, Schetelig J, Kramer M, Schmidt H, Schmidt AH, Thiede C, et al. Prediction of hematopoietic stem cell yield after mobilization with granulocyte-colony-stimulating factor in healthy unrelated donors. Transfusion. 2015;55(12):2855–63.

33. Bailen R, Perez-Corral AM, Pascual C, Kwon M, Serrano D, Gayoso J, et al. Factors predicting peripheral blood progenitor cell mobilization in healthy donors in the era of related alternative donors: Experience from a single center. Journal of clinical apheresis. 2019.

34. Martino M, Callea I, Condemi A, Dattola A, Irrera G, Marcuccio D, et al. Predictive factors that affect the mobilization of CD34(+) cells in healthy donors treated with recombinant granulocyte colony-stimulating factor (G-CSF). Journal of Clinical Apheresis. 2006;21(3):169–75.

35. Richa E, Papari M, Allen J, Martinez G, Wickrema A, Anastasi J, et al. Older age but not donor health impairs allogeneic granulocyte colony-stimulating factor (G-CSF) peripheral blood stem cell mobilization. Biology of Blood & Marrow Transplantation. 2009;15(11):1394–9.

36. Vasu S, Leitman SF, Tisdale JF, Hsieh MM, Childs RW, Barrett AJ, et al. Donor demographic and laboratory predictors of allogeneic peripheral blood stem cell mobilization in an ethnically diverse population. Blood. 2008;112(5):2092–100.

37. Chen SH, Yang SH, Chu SC, Su YC, Chang CY, Chiu YW, et al. The role of donor characteristics and post-granulocyte colony-stimulating factor white blood cell counts in predicting the adverse events and yields of stem cell mobilization. International Journal of Hematology. 2011;93(5):652–9.

38. de Lavallade H, Ladaique P, Lemarie C, Furst S, Faucher C, Blaise D, et al. Older age does not influence allogeneic peripheral blood stem cell mobilization in a donor population of mostly white ethnic origin. Blood. 2009;113(8):1868–9.

39. Furst D, Hauber D, Reinhardt P, Schauwecker P, Bunjes D, Schulz A, et al. Gender, cholinesterase, platelet count and red cell count are main predictors of peripheral blood stem cell mobilization in healthy donors. Vox sanguinis. 2019.

40. Martino M, Gori M, Moscato T, Naso V, Ferreri A, Provenzano F, et al. The challenge to predict mobilized peripheral blood stem cells on the fourth day of G-CSF treatment in healthy donors: the predictive value of basal CD34+ cell and platelet counts. Biology of blood and marrow transplantation : journal of the American Society for Blood and Marrow Transplantation. 2019.

41. Martino M, Gori M, Pitino A, Gentile M, Dattola A, Pontari A, et al. Basal CD34(+) Cell Count Predicts Peripheral Blood Stem Cell Mobilization in Healthy Donors after Administration of Granulocyte Colony-Stimulating Factor: A Longitudinal, Prospective, Observational, Single-Center, Cohort Study. Biol Blood Marrow Transplant. 2017;23(7):1215–20.

42. Wang TF, Liou YS, Chang HH, Yang SH, Li CC, Wang JH, et al. Correlation of Body Mass Index and Proinflammatory Cytokine Levels with Hematopoietic Stem Cell Mobilization. J Clin Med. 2022;11(14).

43. Pruszczyk K, Bartnik K, Bogusz K, Farhan R, Cwil D, Jastrzebska A, et al. Prior blood donations do not affect efficacy of G-CSF mobilization nor outcomes of haematopoietic stem cell collection in healthy donors. Vox sanguinis. 2019.

44. Hsu JW, Wingard JR, Logan BR, Chitphakdithai P, Akpek G, Anderlini P, et al. Race and ethnicity influences collection of granulocyte colony-stimulating factor-mobilized peripheral blood progenitor cells from unrelated donors, a Center for International Blood and Marrow Transplant Research analysis. Biology of Blood & Marrow Transplantation. 2015;21(1):165–71.

45. Lysak D, Hrabetova M, Vrzalova J, Koza V, Navratilova J, Svoboda T, et al. Changes of cytokine levels during granulocyte-colony-stimulating factor stem cell mobilization in healthy donors: association with mobilization efficiency and potential predictive significance. Transfusion. 2011;51(2):319–27.

46. Chen J, Burns KM, Babic A, Carrum G, Kennedy M, Segura FJ, et al. Donor body mass index is an important factor that affects peripheral blood progenitor cell yield in healthy donors after mobilization with granulocyte-colony-stimulating factor. Transfusion. 2014;54(1):203–10.

47. Farhadfar N, Ahn KW, Bo-Subait S, Logan B, Stefanski HE, Hsu JW, et al. The Impact of Pre-Apheresis Health Related Quality of Life on Peripheral Blood Progenitor Cell Yield and Donor’s Health and Outcome: Secondary Analysis of Patient-Reported Outcome Data from the RDSafe and BMT CTN 0201 Clinical Trials. Transplantation and cellular therapy. 2022;28(9):603.e1–.e7.

48. Kroger N, er AR. Dose and schedule effect of G-GSF for stem cell mobilization in healthy donors for allogeneic transplantation. Leukemia & Lymphoma. 2002;43(7):1391–4.

49. Martino M, Moscato T, Barilla S, Dattola A, Pontari A, Fedele R, et al. Mobilization of hematopoietic progenitor stem cells in allogeneic setting with lenograstim by subcutaneous injection, in daily or twice-daily dosing: a single-center prospective study with historical control. Transfusion. 2015;55(8):2032–8.

50. Kroger N, Renges H, Kruger W, Gutensohn K, Loliger C, Carrero I, et al. A randomized comparison of once versus twice daily recombinant human granulocyte colony-stimulating factor (filgrastim) for stem cell mobilization in healthy donors for allogeneic transplantation. British Journal of Haematology. 2000;111(3):761–5.

51. Anderlini P, Donato M, Lauppe MJ, Huh YO, Martin TG, Chan KW, et al. A comparative study of once-daily versus twice-daily filgrastim administration for the mobilization and collection of CD34+ peripheral blood progenitor cells in normal donors. British Journal of Haematology. 2000;109(4):770–2.

52. Pinar IE, Ozkocaman V, Ozkalemkas F, Durgut H, Dakiki B, Ersal T, et al. Is split-dose better than single-dose? Results of Turkish Stem Cell Coordination Center (TURKOK) donors in the era of rising biosimilar G-CSF. Journal of clinical apheresis. 2022;37(5):430–7.

53. Lysák D, Hejretová L, Hrabetová M, Jindra P. Should We Stop Collecting the Preoperative Autologous Blood before Bone Marrow Harvest? Journal of clinical medicine. 2021;10(10).

54. Tomblyn M, Gordon LI, Singhal S, Tallman MS, Williams S, Winter JN, et al. Use of total leukocyte and platelet counts to guide stem cell apheresis in healthy allogeneic donors treated with G-CSF. Bone Marrow Transplantation. 2005;36(8):663–6.

55. Wang TF, Chen SH, Yang SH, Su YC, Chu SC, Li DK. Poor harvest of peripheral blood stem cell in donors with microcytic red blood cells. Transfusion. 2013;53(1):91–5.

56. Pruszczyk K, Plachta M, Urbanowska E, Król M, Król M, Feliksbrot-Bratosiewicz M, et al. Seasonal variation of human physiology does not influence the harvest of peripheral blood CD34+ cells from unrelated hematopoietic stem cell donors. Transfusion and apheresis science : official journal of the World Apheresis Association : official journal of the European Society for Haemapheresis. 2020:102917-.

57. Schulz M, Karpova D, Spohn G, Damert A, Seifried E, Binder V, et al. Variant rs1801157 in the 3’UTR of SDF-1s does not explain variability of healthy-donor G-CSF responsiveness. PLoS ONE [Electronic Resource]. 2015;10(3):e0121859.

58. Myllymaki M, Redd R, Reilly CR, Saber W, Spellman SR, Gibson CJ, et al. Short telomere length predicts nonrelapse mortality after stem cell transplantation for myelodysplastic syndrome. Blood. 2020;136(26):3070–81.

59. Shaw BE, Logan BR, Spellman SR, Marsh SGE, Robinson J, Pidala J, et al. Development of an Unrelated Donor Selection Score Predictive of Survival after HCT: Donor Age Matters Most. Biol Blood Marrow Transplant. 2018;24(5):1049–56.

60. Bilgin YM. Use of Plerixafor for Stem Cell Mobilization in the Setting of Autologous and Allogeneic Stem Cell Transplantations: An Update. Journal of blood medicine. 2021;12:403–12.

61. de Haas M, Kerst JM, van der Schoot CE, Calafat J, Hack CE, Nuijens JH, et al. Granulocyte colony-stimulating factor administration to healthy volunteers: analysis of the immediate activating effects on circulating neutrophils. Blood. 1994;84(11):3885–94.

62. Szmigielska-Kaplon A, Szemraj J, Hamara K, Robak M, Wolska A, Pluta A, et al. Polymorphism of CD44 influences the efficacy of CD34(+) cells mobilization in patients with hematological malignancies. Biol Blood Marrow Transplant. 2014;20(7):986–91.

63. Bogunia-Kubik K, Gieryng A, Gebura K, Lange A. Genetic variant of the G-CSF receptor gene is associated with lower mobilization potential and slower recovery of granulocytes after transplantation of autologous peripheral blood progenitor cells. Cytokine. 2012;60(2):463–7.

64. Camurdanoglu BZ, Esendagli G, Ozdemir E, Canpinar H, Guc D, Kansu E. The effect of granulocyte colony stimulating factor receptor gene missense single nucleotide polymorphisms on peripheral blood stem cell enrichment. Cytokine. 2013;61(2):572–7.

65. Garciaz S, Sfumato P, Granata A, Imbert AM, Fournel C, Calmels B, et al. Analysis of a large single institution cohort of related donors fails to detect a relation between SDF1/CXCR4 or VCAM/VLA4 genetic polymorphisms and the level of hematopoietic progenitor cell mobilization in response to G-CSF. PloS one. 2020;15(3):e0228878-e.

66. Mishima S, Matsuda C, Ishihara T, Nagase M, Taketani T, Nagai A. Single nucleotide polymorphisms of the DGKB and VCAM1 genes are associated with granulocyte colony stimulating factor-mediated peripheral blood stem cell mobilization. Transfus Apher Sci. 2017;56(2):154–9.

67. Benboubker L, Watier H, Carion A, Georget MT, Desbois I, Colombat P, et al. Association between the SDF1-3’A allele and high levels of CD34(+) progenitor cells mobilized into peripheral blood in humans. Br J Haematol. 2001;113(1):247–50.

68. Xiang J, Shi M, Fiala MA, Gao F, Rettig MP, Uy GL, et al. Machine learning-based scoring models to predict hematopoietic stem cell mobilization in allogeneic donors. Blood Adv. 2022;6(7):1991–2000.

